# Childhood features associated with men’s Beliefs in God and/or a Divine Power in England

**DOI:** 10.1101/2025.07.11.25331346

**Authors:** Jean Golding, Jimmy Morgan, Steven Gregory, Yasmin Iles-Caven, Alan Emond, Kate Northstone, Crystal Park

## Abstract

Research has linked childhood experiences with subsequent religiousness and/or spirituality, but this work generally focuses on general religiousness or worship service attendance. Very little is known about how childhood experiences influence individuals’ religious or spiritual beliefs (RSBs). We have investigated whether the childhoods of men who have such a belief (measured by the question ‘‘Do you believe in God or in some Divine power?’) differ from those of their peers without such a belief. We used data collected from partners of pregnant women with expected dates of delivery in 1991-2 residing in a specified area of south-west England. These men answered detailed questionnaires about their childhood. Of the 213 descriptors of their childhood, 48 were associated with RSBs at P<0.01, 23 times the number that would be expected by chance. A series of stepwise logistic regression analyses were carried out to identify the key independent variables that would need to be considered in studies assessing whether the RSB of these men was associated with their subsequent health. We found that the childhood features that distinguished the men with this belief from their peers fell into three groups: (i) indicators of a happy secure childhood; (ii) severe trauma (such as being abused or being seriously ill), and (iii) less acute childhood circumstances (such as their criminal behaviour, being expelled from school and having parents going through a divorce). The items in the first two of these groups were associated with increased likelihood of RSB, whereas the items in (iii) were associated with a reduced likelihood. These results need to be repeated elsewhere with different population groups before they can be generalised, but meanwhile they will form a basis for the choice of confounders, mediators or moderators in studies of the health and well-being of these men.

## Background

Religious/spiritual beliefs (RSBs) are generally recognized as a core dimension of religiousness, yet surprisingly little research has explicitly focused on the roles that they play in health, well-being, and behaviour [1,2]. This lack of attention contrasts with the focus on demonstrable factors such as attendance at a place of worship [3], although, in Britain and many other places, many more individuals have RSBs than regularly attend a place of worship [4]. As research increasingly concerns the ways in which RSBs may be associated with health [3,5], it is important to understand the characteristics leading to these beliefs, so that they can be considered in any analyses since these associated factors may act as confounders, moderators or mediators.

Research on the childhood factors associated with religiosity in general have included minority ethnicity, peers’ church attendance during high school, attendance at a faith school, maternal religiosity, church attendance during childhood, and the importance mothers place on childhood religious training [6–8]. There is some suggestion that stressful and traumatic life events in childhood can influence adult religiousness although findings are contradictory. Leo and colleagues [9] carried out a literature review and concluded that most people do not change their religious beliefs after a trauma, but significant changes occur for a small proportion of people - either increasing or decreasing their religious beliefs.

Although, the so-called ACEs (Adverse Childhood Experiences) have been associated with increased risk of adult outcomes as varied as ill health and mortality, alcoholism and depression [10–14], evidence is missing as to whether the RSB of the individual plays any part in these associations. The present research therefore investigates the possible contribution of childhood histories (including positive as well as negative features) to RSB among adult men (we have not included women in this paper as their experiences during childhood differ considerably from those of men; they will be considered in a subsequent study). We have used data collected by the longitudinal pre-birth cohort – the Avon Longitudinal Study of Parents and Children (ALSPAC) for this project, and identified those with a positive RSB as those answering ‘yes’ to the question: ‘Do you Believe in God or some Divine Power?’

The aim of this paper is to investigate a broad array of potential childhood influences on the RSBs of men and deciphering which variables are independently associated. There are several reasons for looking at this question. First and foremost is sheer scientific curiosity. Second is the practical question concerning our proposed future studies of the relationship between RSB and health, as to which variables should be considered as possible statistical confounders, mediators or moderators. Third is to identify a base from which to compare the independent childhood factors associated with RSB in adult men in ALSPAC (almost all of whom are fathers) with other groups of individuals, including women and their offspring in adolescence and in childhood. Such future analyses will allow an insight as to whether any associations shown here are consistent between the sexes and across generations.

Here we have included approximately 10,000 men who were the partners of pregnant women in 1990-2. Before their child was born the men were asked whether they believed in God or a Divine Power (RSB). The investigation is hypothesis-free and uses an exposome approach [15]. Exposomes are used to test associations with environmental exposures, whether physical, biological, psychological or psychosocial.

## Material and Methods

### The ALSPAC study

In 1990, in the English county of Avon, the Avon Longitudinal Study of Parents and Children (ALSPAC), a pre-pregnancy longitudinal study, began with the aim of identifying the factors, (both environmental and genetic) that influence a child’s health and well-being [16]. To this end, the study was designed to enrol all pregnant women resident in the defined area with an expected date of delivery between 1^st^ April 1991 and 31^st^ December 1992. Approximately 75-80% (n = 14,541) of eligible women joined the study [17,18]. They were asked to invite their partners to take part but were under no obligation to do so [19,20].

Data were collected using self-completed questionnaires, posted directly to the mothers. A questionnaire for her partner (with a reply-paid envelope) was sent to the mother to hand to her partner if she so wished. This arrangement was made on the advice of the study’s Ethics Committee (see [19] for further details of the discussions and decisions made). Thus, the decision as to whether the mothers’ partner took part in the study was predominately up to the study mother. This obviously had the problem of a larger drop-out rate than found for the mothers themselves. Details of the attrition rate at various times are given elsewhere [20]. For our analyses we selected the 9774 men who answered the initial question on RSB during their partner’s pregnancy. They had a mean age of 30.6 (S.D. 5.8) years; 20% had been educated to university degree level, 76% were living in property that they either owned or were purchasing using a mortgage. Just 3% belonged to a non-White minority ethnic group.

### Ethics Approval and Consent to Participate

Ethical approval for the study was obtained from the ALSPAC Ethics and Law Committee and the Local Research Ethics Committees. Implied consent from participants for the use of data collected via questionnaires and clinics was assumed following the recommendations of the ALSPAC Ethics and Law Committee at the time [19]. ALEC has remained independent of ALSPAC throughout and has been approved by the American Institutional Review Board (IRB no.00003312). The submission of a completed questionnaire, either on paper or online, was considered to be written consent from participants to use their data for research purposes. Study participants have the right to withdraw their consent for elements of the study or from the study entirely at any time. Full details of the ALSPAC consent procedures are available on the study website (http://www.bristol.ac.uk/alspac/researchers/research-ethics/). There is a strict ethical divide in that identifying information is only available to very few staff for the purposes of questionnaire or face-to-face clinic administration. No researcher has access to identifying information. Data reported in this paper were initially accessed 1^st^ September, 2021.

### The outcome

During pregnancy each parent-to-be was asked questions concerning religious and spiritual beliefs and behaviours (see [21]) for further details]. The first such question was: ‘Do you believe in God or in some Divine power?’ with possible responses: yes/not sure/no. For the analyses in this paper, we compare those who answered ‘yes’ with the rest.

### The exposures

Information on the men’s childhood was collected during pregnancy from the men themselves using two self-completion questionnaires and a further one at 33 months after the birth of the ALSPAC study child. We selected for this exposome all the variables collected that related to his background and events during his childhood prior to the age of 17 years and where the amount of information was sufficient for analysis (in general, the proportion with a positive answer to a particular question was >2% of those asked). This resulted in 213 variables which we initially divided into five groups, three of which were focussed on specific ages: (i) the first 5 years; (ii) pre-puberty (6-11years); and (iii) adolescence (12-16 years); group (iv) comprised life events occurring < 17 years but without a specific age stated, and group (v) comprised information that was collected later (33 months after the birth of the child) on the childhood environment (including home/parental ethos, attitudes to school and his criminal behaviours) (see SFig). The fifth group was treated separately as the numbers of men replying to these questions were substantially less than those answering in pregnancy (∼ 5000 compared with ∼9500). Further information on the actual questions used can be obtained by reading the descriptions of the variables and coding strategy used in the ‘built file’ section of the ALSPAC Data Dictionary for the PA, PB and PF questionnaires.

Please also note that the study website contains details of all the data that are available through a fully searchable data dictionary and variable search tool: http://www.bristol.ac.uk/alspac/researchers/our-data/

The five groups of variables were themselves subdivided into categories as shown in Figure 1, resulting in a total of 18 different strata. Of the 213 variables involved, there were several that were similar to others in a different stratum. This was one of the factors that drove the way in which the statistical methods were applied.

**Figure 1.**
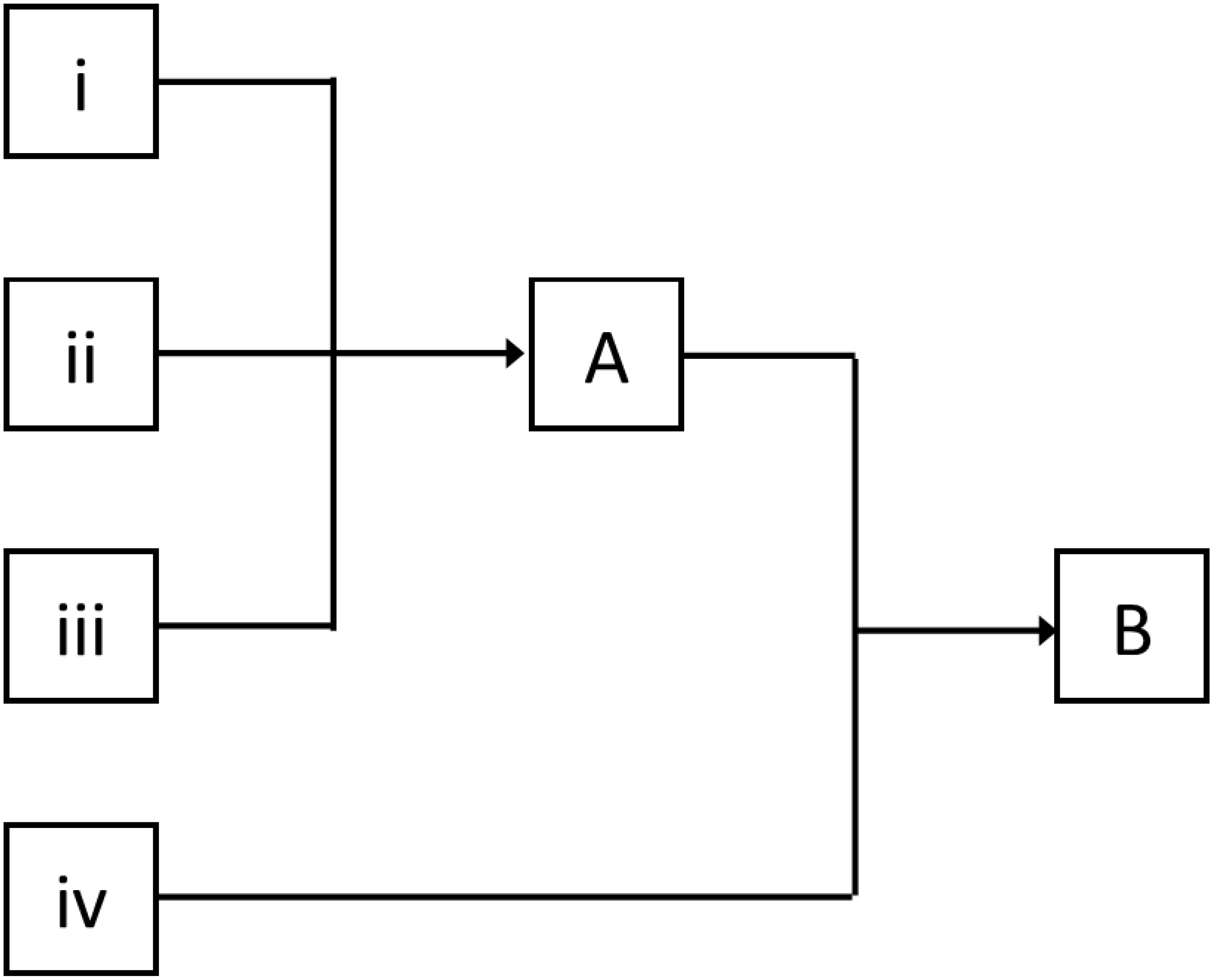
The analysis plan

### The Statistical Methodology

The identification of independent factors associated with the men’s RSB at the time of pregnancy used an exposome technique as developed earlier [22, 23]. In general, this involved: (1) an unadjusted analysis of each factor by the outcome and recording the relevant P-value. (2) Within each group identifying all P values that were associated with RSB in unadjusted analyses and determining whether there were more associations at P<0.05, 0.01, 0.001 and 0.0001 than would be expected by chance if the factors were independent of one another (this is roughly analogous to a Q-Q probability plot as often used in GWAS analyses. (3) For groups in which there were substantially more associations at P<0.01 than expected by chance, carrying out the same strategy for each subgroup and determining, for each, the numbers observed and expected at the same P values as above. (4) Using, within each selected group or groups, a backward stepwise logistic regression to select those variables independently associated with RSB. (5) Combining the variables identified as independent from each group with those identified in this way from other groups sequentially. (6) using a final stepwise logistic regression analysis to identify independent variables to be included in a final model.

Because the numbers of individuals available for analysis were consistently greater for the first four groups, these were first analysed together to create Model A. The independent items from Group (v) were then added to those in Model A and stepwise regression identified the final variables (Model B).

For each analysis we have documented the numbers of items on which the models were derived, the numbers of variables involved and a measure of goodness-of-fit (GoF) based on the pseudo-R^2^ so that the interested researcher can decide which combination might be best used as confounders, mediators or moderators when analysing RSB, based on the statistical power and/or hypothesis being tested.

## Results

Of the 9774 men who answered the question on Belief in God or a Divine Power, 25% stated that they did have such a belief. This proportion was compared between different aspects of the groups and subgroups outlined in Figure 1. The overall distribution of P values for the unadjusted associations for each group of variables is shown in Table 1 and compared with the numbers that would be expected by chance. Overall, there were 6 times more associations than expected at P<0.05, 23 times more at P<0.01, 150 times more at P<0.001 and 1100 times more likely at P<0.0001.

**Table 1.**
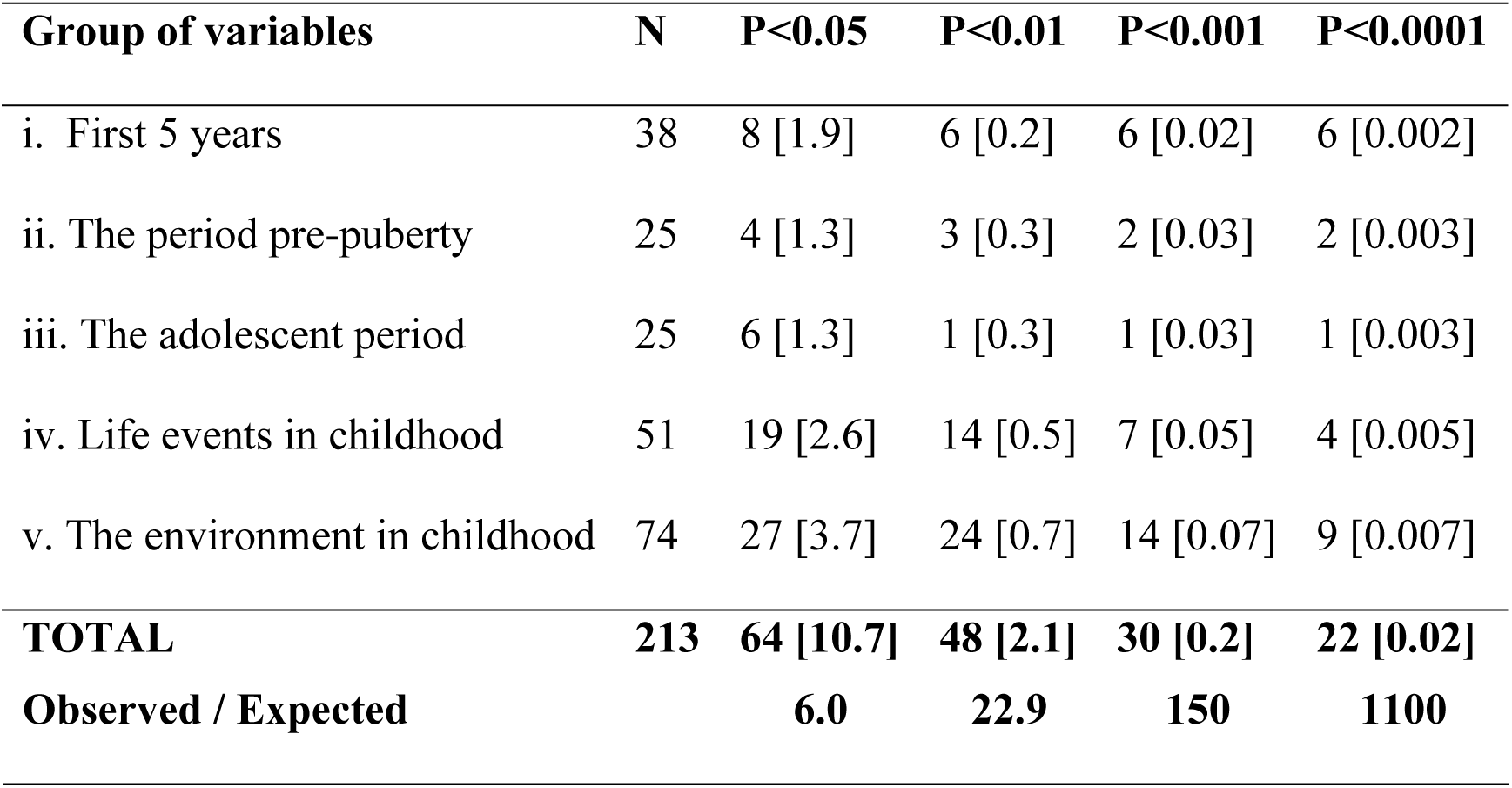
The numbers of variables considered, and the numbers associated with having an RSB at various P values; in square brackets are the numbers expected.

**Table 2.**
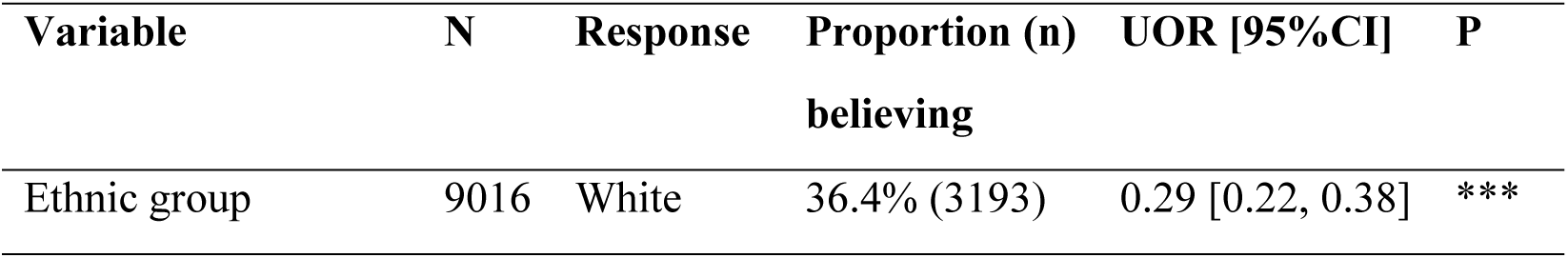
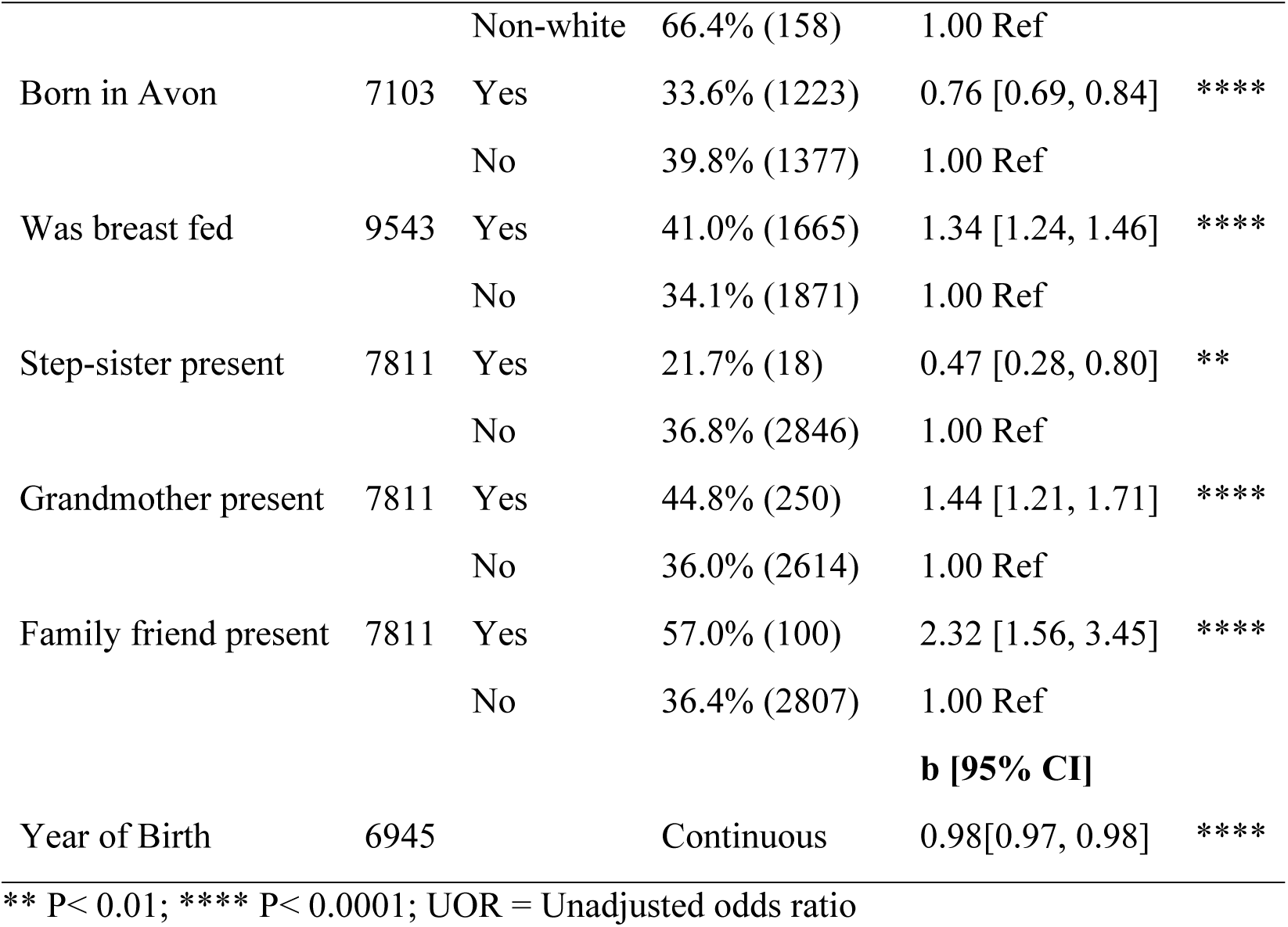
Variables in groups (i) and (ii): related to the men’s RSB at P<0.01. Data shown are the proportions who are believers, and the unadjusted odds ratio related to each response, with 95% confidence intervals (CI); year of birth is shown with its regression coefficient.

The ways in which the different subgroups contributed to this overall distribution of P-values is shown in Table S1. This indicates that the following categories were of no further interest to the analysis as they had no more than the expected distribution of P values: accidental injuries at each of the three age groups; the household composition in early childhood and in adolescence; life events at specified age groups; neglect and abuse when asked at 33 months after the pregnancy; and criminal behaviour in pre-teenage childhood. The remainder for further analyses were: measures at birth or in infancy; features of the household when he was aged 6-11 years; ‘other’ features of his adolescence (which comprised: smoking cigarettes at this time; childhood life events (items of scale); schooling and home environment; psychosocial background; and criminal behaviour when a teenager.

To simplify the subsequent analyses, the variables with more associations than expected at P<0.01 were reorganised into four groups: (i) the first years; (ii) puberty and adolescence; (iii) life events; and (iv) the psychosocial environment in his childhood (a group of variables asked about the family dynamics 33 months after the birth). First, we analysed groups (i) – (iii) to create Model A and considered (iv) separately because the numbers answering these questions were smaller than for the other groups, before combining them with A to form Model B (Figure 1).

### (i) The first 11 years

The variables grouped as relative to the first 11 years of the mens’ lives showed substantially more associations at P<0.01 than were expected by chance. These comprised ethnic group (white /non-white); whether born in Avon; whether he had been breast fed; and whether a grandmother, step-sister or family friend had been present in the household when he was aged 6-11 years. The unadjusted analyses indicated that low levels of belief were apparent if he was white, had been born in Avon or there was a step-sister present, whereas higher than expected odds of RSB were apparent if he had been breast fed, and either his grandmother or a family friend had been resident in the household (Table S2). His year of birth was such that the more recently he had been born (i.e. the younger he was), the less likely was he to believe. These variables were offered to stepwise regression, but all remained in the analysis, implying that all were statistically independent of one another.

### (ii) Puberty and Adolescence

For this section we combined the other variables at P<0.01 which occurred in the sections: Pre-puberty and Adolescence and Schooling and Home environment from the Environment group. After excluding those variables already considered, there were 10 variables which unadjusted analyses had identified at P<0.01; of these the following were associated with reduced likelihood of subsequent RSB: smoking in adolescence; being expelled from school; and having truanted at age 11 or later. In contrast, the following were associated with increased likelihood of having an RSB: he was happy at ages 6-11 years; his father was present in the household during his adolescence; he failed an important exam; he found school a valuable experience; he was absent from school due to illness <11 years; he reported that his mother was very caring, but also over-protective (Table S3). When these 11 variables were offered to stepwise logistic regression, two items dropped out: the father’s presence in the home and being expelled from school.

### (iii) Life events during childhood

A series of 31 items were included in the childhood life event questions derived from Coddington [24]. Nine of these, together with the weighted life events score were associated with RSB at P<0.01. Of these, those items predictive of lower likelihood of RSB were: parents separating; parents divorced; a parent remarrying; and being in trouble with the police. Increased likelihood of RSB was found if there was a childhood history of a friend dying; his being in hospital; having had a serious accident; a parent being emotionally cruel to him; and being sexually abused. Although there was no difference between the men who were believers and the non-believers in the actual numbers of life events they had experienced, there was a strong difference in their life event scores weighted by the degree to which they stated that they had been affected by the event in that those who were believers subsequently reported being more affected (Table S4).

When these 10 variables were offered to stepwise regression, six of the items dropped out: death of a friend, hospital admission, serious accident, separation of parents, parent remarrying, and a parent being emotionally cruel. Thus, the only variables remaining in this group were parents having divorced, his being sexually abused, in trouble with the police and the weighted childhood life events score.

### Model A: Combining (i), (ii) and (iii)

In Table 3 can be seen the final variables independently associated with each of groups (i), (ii) and (iii). All were offered to a further backwards logistic regression to form Model A. Of the variables in group (i), two dropped out: the presence of a step-sister and the man’s year of birth. Of the 9 variables remaining in group (ii) three dropped out – smoking in adolescence, failing an important exam and truanting at age 11+. Finally, of the four variables left in (iii), none were eliminated at this stage. Thus, Model A comprises 15 variables independently associated with RSB.

**Table 3.** Adjusted associations with RSB: (OR [95%CI]) for binary variables^a^ and linear coefficients [95% CI] for continuous variables^b^, between the four groups of childhood variables depicted in Tables S1-S3, using backward logistic regression.

| Variable | Group (i) | Group (ii) | Group (iii) | Model A |
| --- | --- | --- | --- | --- |
| Ethnic group is white<sup>a</sup> | 0.56<br>[0.37, 0.85] |  |  | 0.45<br>[0.38, 0.67] |
| Born in Avon<sup>a</sup> | 0.83<br>[0.74, 0.93] |  |  | 0.84<br>[0.80,1.00] |
| Was breast fed<sup>a</sup> | 1.25<br>[1.12, 1.40] |  |  | 1.21<br>[1.08,1.35] |
| Step-sister was present in household<sup>a</sup> | 0.46<br>[0.23, 0.89] |  |  |  |
| Grandmother present in household<sup>a</sup> | 1.36<br>[1.19, 1.67] |  |  | 1.23<br>[1.05,1.57] |
| Family friend present in household<sup>a</sup> | 3.10<br>[1.86, 5.19] |  |  | 2.44<br>[1.50,3.96] |
| Year of birth<sup>b</sup> | 0.99<br>[0.98, 0.99] |  |  |  |
| Very happy aged 6-11<sup>a</sup> |  | 1.15<br>[1.02, 1.29] |  | 1.19<br>[1.05,1.36] |
| Started smoking < 16 years<sup>a</sup> |  | 0.79<br>[0.65, 0.95] |  |  |
| Failed an important exam<sup>a</sup> |  | 1.18<br>[1.05, 1.34] |  |  |
| Always liked school<sup>a</sup> |  | 1.32<br>[1.10, 1.58] |  | 1.22<br>[1.01,1.48] |
| Felt school was a valuable experience<sup>a</sup> |  | 1.21<br>[1.12, 1.31] |  | 1.14<br>[1.04, 1.26] |
| Truanted aged 11+<sup>a</sup> |  | 0.79<br>[0.66, 0.95] |  |  |
| Away from school through illness <11<sup>a</sup> |  | 1.40<br>[1.10, 1.78] |  | 1.40<br>[1.08,1.80] |
| Maternal care score<sup>b</sup> |  | 1.02<br>[1.00, 1.03] |  | 1.02<br>[1.00,1.03] |
| Maternal overprotection score<sup>b</sup> |  | 1.04<br>[1.03, 1.06] |  | 1.04<br>[1.02, 1.05] |
| Parents divorced<sup>a</sup> |  |  | 0.63<br>[0.55, 0.73] | 0.71<br>[0.58,0.86] |
| Was sexually abused<sup>a</sup> |  |  | 1.58<br>[1.09, 2.28] | 2.02<br>[1.24,3.30] |
| In trouble with the police<sup>a</sup> |  |  | 0.65<br>[0.58, 0.73] | 0.69<br>[0.59,0.80] |
| Weighted life events score<sup>b</sup> |  |  | 1.02<br>[1.02, 1.03] | 1.02<br>[1.01,1.03] |
| **Number in model** | **5468** | **6343** | **9758** | **5809** |

CI = Confidence Interval; <sup>a</sup>Binary variable; <sup>b</sup>Continuous variable

### (iv) Psychosocial variables

Seven psychosocial variables showed unadjusted associations at P<0.01. Of these, six were associated with an increased likelihood of having RSB: having a family that did things together; parents who were affectionate; had a relationship that was frightening, friendly or respectful. The only item associated with a lower likelihood of RSB concerned whether the parents’ relationship was remote (Table S5). When these six variables were included in a stepwise regression, just three variables remained: whether the parents’ relationship was frightening, affectionate or respectful (see column iv in Table 3).

### Should the final model be A or B?

Combining the variables from the stepwise analysis of (iv) with those in Model A, and carrying out a further stepwise regression, six variables dropped out, leaving 13 variables in Model B (Table 3). This compares with 15 variables in Model A and raises the question as to which is the most appropriate to consider when choosing potential confounders for analyses involving RSB.

In Table 4, the characteristics of each of the independent sets of variables from each of (i) – (iv) and Models A and B are compared regarding the number of variables in the model, the number of records used, and the goodness of fit (GoF) as measured using the pseudo-R^2^. The advantage of using Model B is that there are slightly fewer variables included, and the GF is slightly greater. The disadvantage is that the number of records available is considerably smaller (by approximately 2500 records) in Model B. Thus, for analyses using the identified factors as mediators, moderators or potential confounders, the items in Model A would allow for a more powerful analysis than if Model B items were used because of the increased number of cases available.

**Table 4.** Characteristics of each backwards logistic regression analysis after elimination of factors that did not remain in the model; outcome = RSB.

| Model | N | GF | No. variables in model |
| --- | --- | --- | --- |
| (i) | 5468 | 1.29 | 7 |
| (ii) | 6343 | 1.77 | 9 |
| (iii) | 9758 | 0.98 | 4 |
| (iv) | 4382 | 0.77 | 4 |
| Model A | 5809 | 3.03 | 15 |
| Model B | 3325 | 3.14 | 13 |

GF = Goodness of fit measure

## Discussion

This set of analyses was undertaken with the assumption that features of individuals’ childhoods may influence their subsequent RSBs. Within this concept, we have conducted a hypothesis-free set of analyses concerning the childhood features that were independently related to whether a man was likely to have such a belief at a time when he was about to be a father. We have shown that the features associated with a reduction in his odds of having RSB comprised whether he was White, whether his parents had divorced during his childhood and whether he had been in trouble with the police as a teenager (Models A and B). Increased likelihood of RSB occurred when exposed to warm positive childhoods, especially if a family friend had been present in the household, whether he was happy during his pre-puberty years, he felt school was a valuable experience and his parents were affectionate and respectful of one another. Nevertheless, his mother was more likely to be rated by him as having been over-protective. In contrast he was also more likely to have an RSB if there had been traumas in his childhood such as: (a) a serious illness before he was 11; (b) being sexually abused; (c) his parents having a relationship which he found frightening. An additional example of increased trauma in childhood among these men concerned the fact that those who subsequently had an RSB had rated the life events they experienced as affecting them more than those who had not had an RSB.

The fact that adverse occurrences were associated with a reduced likelihood of having an RSB was not surprising [9], nor was the likelihood of an increased odds of having RSB when the child was born into a warm affectionate family. However, the associations between increased likelihood of belief when there had been traumatic events in childhood such as a serious illness, being abused, having frightening parents and their ratings of other events as affecting them more strongly highlights the complexity of links between highly stressful events and religious or spiritual beliefs [9]. Unfortunately, we are unable to assess temporality—did belief pre-date or follow the trauma – since the retrospective recording of the traumatic events occurred at almost the same time as the recording of the RSBs. There is scant recent literature on this subject, although Ryan [25] in reviewing the literature, noted that survivors of childhood violence often mentioned the importance of spirituality in their survival and recovery. She also noted, however, that the effects of childhood traumas on spirituality can either be enhancing or damaging. From the results here it would also appear that the type of trauma is a key factor in whether the result is an increase or decrease in RSB.

Among the non-traumatic variables that entered both Models A and B was an increased incidence of RSB among men who had been breast fed. This could indicate a positive bond between the man and his mother, or it may have biological significance. It has been shown that breast-fed individuals have higher levels of cognition, and that this was true of the 1940s in the UK when it was mainly the uneducated classes that breastfed, and of the more recent cohorts where it was the educated classes that breastfed. The question was answered convincingly with numerous observational studies and even a randomised trial [26, 27]. The component of breast milk that was not found in cows’ milk substitutes at the time the men were born was the omega-3 fatty acid docosahexaenoic acid (DHA), which is known to impact the cognitive ability of the child [28]. This raises the question as to whether DHA might influence the development of the brain in ways that promote beliefs including religious ones. Some slight corroborating evidence to support this lies in the fact that we have shown that adults with an RSB are more likely to have a diet that includes fish, which is the main dietary source of (DHA) [29].

The analyses here rigorously excluded those items that did not remain in the stepwise regression. The items that failed to meet the P<0.01 requirement were eliminated – but this P value is an arbitrary choice, chosen to reduce the likelihood of including exposures that were not valid predictors. However, that does not necessarily mean that they would not have been valid components of the model. It is also true that there may well be other features of childhood that were predictive of an RSB of which we had no knowledge; these would have contributed to possible residual confounding.

### Strengths and Limitations

The major strength of this study comprises the sample definition, which included a large proportion of men who were enrolled during the pregnancy of their female partner with no bias in terms of their religion or childhood background. Although these men were assumed to be the biological fathers of the study children, this was not necessarily so, although they usually took the role of father within the household. A second advantage derives from the fact that the questions enquiring about their childhood were distinct from those concerning their RSBs, and there was no suggestion that there was any link expected between the two.

Thirdly, there was a comprehensive wealth of information on the childhoods of these individuals. A future strength lies in the fact that this is a longitudinal study, and these men are being followed up into the future. Finally, the focus on RSB is a strength of the study in that very little research has documented the childhood precursors of adults’ belief in God/the divine, so this research builds on previous studies of general religiousness or religious behaviour.

There are, however, a number of limitations including: (a) the omission from the study of men who were single, infertile or did not want children; (b) the variables used to document the men’s childhoods may have omitted important features that may have changed the results if included (e.g. whether they were members of a religious family or attended a faith school); (c) because of the nature of the population at the time, the majority of the men involved had a Christian background, even if they were agnostic or atheist at the time of the survey - consequently extrapolation from these results to religious or spiritual beliefs in general may not be relevant; (d) information on their childhoods was gained retrospectively – this may have resulted in some distortion of memory, it is probably true that the most severely traumatic events will be remembered (however, see the comments of Maughan [30]). Despite these limitations, we suggest that the results from this study will provide a useful comparison with other studies in the literature.

## Conclusions

The events occurring within the childhoods of the men in this study showed associations that can be summarised as: (i) certain adverse events in childhood were associated with a positive RSB; (ii) other adverse events concerning the behaviour of the child (e.g. criminality, expulsion from school) were associated with being less likely to have RSB; and (iii) evidence of a warm loving home was associated with an increased likelihood of RSB. However, it is important to ensure that comparable analyses are undertaken with different populations before these results can be generalised.

## Author contributions

This publication is the work of the authors; Steven Gregory & Jean Golding will serve as guarantors for the contents of this paper. SG carried out the statistical analyses; JG derived the concept and design of the study, JG, YIC and KN were responsible for funding acquisition and all authors contributed to writing and rewriting of several versions of the paper.

## Conflicts of Interest

No competing interests were disclosed.

## Grant Information

The UK Medical Research Council and Wellcome (Grant ref: 217065/Z/19/Z) and the University of Bristol currently provide core support for ALSPAC. A comprehensive list of grant funding is available on the ALSPAC website (http://www.bristol.ac.uk/alspac/external/documents/grant-acknowledgements.pdf). This research was made possible through the support of a grant from the John Templeton Foundation (61917). The opinions expressed in this publication are those of the authors and do not necessarily reflect the views of the John Templeton Foundation.

## Data Availability

ALSPAC data is available to researchers for particular projects, provided no attempt is made to reveal the identities of the subjects. Guidelines for access are found on the ALSPAC website: www.bristol.ac.uk/alspac/researchers

## Acknowledgements

We are extremely grateful to all the families who took part in this study, the midwives for their help in recruiting them, and the whole ALSPAC team, which includes interviewers, computer and laboratory technicians, clerical workers, research scientists, volunteers, managers, receptionists and nurses.

## Supplementary Tables

**Table S1.** The numbers of variables considered, and the numbers associated with the father having an RSB at various P values in the subgroups of Table 1.

**Table S2.** Pre-adolescent variables in groups (i) and (ii) related to the men’s RSB at P<0.01. Data shown are the proportions who are believers and the unadjusted odds ratio related to each response, with 95% confidence intervals (CI); year of birth is shown with its regression coefficient.

**Table S3.** Variables (ii) related to the father’s later childhood, school and home environments. Data shown are the proportions who are believers and the unadjusted odds ratio relating to each response, with 95% confidence intervals (CI)

**Table S4.** Life events (iii) occurring during fathers’ childhood (<17 years). Data shown are the proportions who are believers and the unadjusted odds ratio relating to each response, with 95% confidence intervals (CI)

**Table S5.** Variables (v) related to his home life during childhood. Data shown are the proportions who are believers and the unadjusted odds ratio relating to each response, with 95% confidence intervals (CI)

## Notes

### Competing Interest Statement

The authors have declared no competing interest.

### Author Declarations

Ethical approval for the study was obtained from the ALSPAC Ethics and Law Committee and the Local Research Ethics Committees. Implied consent from participants for the use of data collected via questionnaires and clinics was assumed following the recommendations of the ALSPAC Ethics and Law Committee (ALEC) at the time [13]. ALEC has remained independent of ALSPAC throughout and has been approved by the American Institutional Review Board (IRB no.00003312). Detailed information on the ways in which confidentiality of the cohort is maintained may be found on the study website: http://www.bristol.ac.uk/alspac/researchers/research-ethics/

## References

1. Park C. Finally, some well-deserved attention to the long neglected dimension of religious beliefs: suggestions for greater understanding and future research. RBB. 2020;10(2):191–197

2. Park CL, Slattery JM. Religiousness and spirituality in health psychology. InAPA handbook of health psychology, Volume 1: Foundations and context of health psychology, 2025 (pp. 569-585). American Psychological Association.

3. Koenig HG, VanderWeele TJ, Peteet JR. Handbook of religion and health. 2024:15–29.

4. Voas D, Bruce S. In Curtice, J., Clery, E., Perry, J., Phillips M. and Rahim, N. (eds.) (2019), British Social Attitudes: The 36th Report, London: The National Centre for Social Research © The National Centre for Social Research. https://natcen.ac.uk/sites/default/files/2023-08/BSA_36.pdf

5. Koenig HG. Suicidal behaviors in diabetics, mental health of cancer patients following curative treatment, substance use disorder in hospitalized psychiatric patients in Botswana, nutritional factors as predictors and mediators of mental health problems in chronic illness, and more. Int J Psych Med. 2023;58:299–301.

6. Gunnoe, M. L., & Moore, K. A. Predictors of religiosity among youth aged 17–22: A longitudinal study of the National Survey of Children. JSSR. 2002;41(4): 613–622.

7. Hardie, J H, Pearce, L D, & Denton, M L The dynamics and correlates of religious service attendance in adolescence. Youth Soc. 2016;48(2):151–175.

8. Hardy, S A, White, J A, Zhang, Z, & Ruchty, J. Parenting and the socialization of religiousness and spirituality. Psychol Relig Spiritual. 2011; 3(3):217–230.

9. Leo, D, Izadikhah, Z, Fein, E C, & Forooshani, S A. The effect of trauma on religious beliefs: A structured literature review and meta-analysis. TVA. 2021;22:161–175.

10. Felitti VJ, Anda RF, Nordenberg D, et al.: Relationship of childhood abuse and household dysfunction to many of the leading causes of death in adults. The Adverse Childhood Experiences (ACE) Study. Am J Prev Med. 1998;14(4): 245–58.

11. Howe LD, Tilling K, Lawlor DA. Studying the life course health consequences of childhood adversity: challenges and opportunities. Circulation. 2015;131(19): 1645–7

12. Hughes K, Lowey H, Quigg Z, et al. Relationships between adverse childhood experiences and adult mental well-being: results from an English national household survey. BMC Pub Health. 2016;16:222.

13. Hughes K, Bellis MA, Hardcastle KA, et al. The effect of multiple adverse childhood experiences on health: a systematic review and meta-analysis. Lancet Pub Health. 2017;2(8):e356–66.

14. Leung JP, Britton A, Bell S. Adverse Childhood Experiences and Alcohol Consumption in Midlife and Early Old-Age. Alcohol Alcohol.2016;51(3):331–8.

15. Vineis P, Robinson O, Chadeau-Hyam M, Dehghan A, Mudway I, Dagnino S. What is new in the exposome? Environ Int. 2020;143:105887.

16. Golding J, Pembrey M, Jones R, et al. ALSPAC--the Avon Longitudinal Study of Parents and Children. I. Study Methodology. Paediatr Perinat Epidemiol. 2001;15(1):74–87.

17. Boyd A, Golding J, Macleod J, Lawlor DA, Fraser A, Henderson J, Molloy L, et al. Cohort Profile: The ‘Children of the 90s’; the index offspring of The Avon Longitudinal Study of Parents and Children (ALSPAC). Int J Epidemiol. 2013; 42:111–127.

18. Fraser A, Macdonald-Wallis C, Tilling K, Boyd A, Golding J, Davey Smith G, Henderson J, et al. Cohort Profile: The Avon Longitudinal Study of Parents and Children: ALSPAC mothers cohort. Int J Epidemiol 2013;42:97–110.

19. Birmingham K: Pioneering ethics in longitudinal study: The early development of the ALSPAC Ethics & Law Committee. Bristol: Policy Press, 2018.

20. Northstone K, Ben Shlomo Y, Teyhan A et al. The Avon Longitudinal Study of Parents and children ALSPAC G0 Partners: A cohort profile. Wellcome Open Res 2023;8:37 (10.12688/wellcomeopenres.18782.1)

21. Iles-Caven Y, Gregory S, Northstone K and Golding J. Longitudinal data on parental religious behaviour and beliefs from the Avon Longitudinal Study of Parents and Children (ALSPAC) Wellcome Open Res 2019;4:38

22. Golding J, Gregory S, Iles-Caven Y, et al. Parental, prenatal, and neonatal associations with ball skills at age 8 using an exposome approach. J Child Neurol. 2014;29:1390–1398.

23. Golding J, Gregory S, Clark R, Ellis G, Iles-Caven Y, Northstone K. Associations between paracetamol (acetaminophen) intake between 18 and 32 weeks gestation and neurocognitive outcomes in the child: A longitudinal cohort study. Paed and Perinat Epidemiol. 2020;34(3):257–66.

24. Coddington RD: The significance of life events as etiologic factors in the diseases of children—II a study of a normal population. J Psychosom Res. 1972;16(3):205–213.

25. Ryan PL. Spirituality among adult survivors of childhood violence: A literature review. J. Transpers. Psychol. 1998;30(1):39.

26. Schack-Nielsen L, Michaelsen KF. Breast feeding and future health. Curr Opin Clin Nutr Metab. Care. 2006; 9(3):289–96.

27. Smithers LG, Kramer MS, Lynch JW. Effects of breastfeeding on obesity and intelligence: causal insights from different study designs. JAMA Pediatr. 2015;169 (8):707–8.

28. Valenzuela A, Nieto MS. Docosahexaenoic acid (DHA) in fetal development and in infant nutrition. Rev. Med. Chile. 2001;129(10):1203–11.

29. Major-Smith D, Morgan J, Emmett P, Golding J, Northstone K. Associations between religious/spiritual beliefs and behaviours and dietary patterns: analysis of the parental generation in a prospective cohort study (ALSPAC) in Southwest England. OSF Preprints 2022

30. Maughan B. Peer Review Report For: Traumatic childhood events of parents enrolled in the Avon Longitudinal Study of Parents and Children (ALSPAC). Wellcome Open Res. 2020;5:65 (10.21956/wellcomeopenres.17331.r38394)

